# Using the Health Belief Model to design a questionnaire aimed at measuring people’s perceptions regarding COVID-19 immunity certificates

**DOI:** 10.1101/2021.11.12.21266257

**Authors:** Corina-Elena Niculaescu, Isabel Karen Sassoon, Irma Cecilia Landa-Avila, Ozlem Colak, Gyuchan Thomas Jun, Panagiotis Balatsoukas

## Abstract

The present short communication paper describes the methodological approach of applying the Health Belief Model to the use COVID-19 immunity certificates in the UK. We designed an online survey including an adaptation of the following Health Belief Model constructs: perceived COVID-19 susceptibility, perceived COVID-19 severity, perceived benefits of using immunity certificates, perceived barriers from using immunity certificates, perceived severity of not using immunity certificates, and perceived vaccination views. The online cross-sectional survey conducted on the 3^rd^ of August 2021 gathered responses from 534 participants aged 18 and older, representative of the UK population in terms of gender, age, and ethnicity.

---

The Health Belief Model (HBM) is a socio-psychological theoretical model developed in the 1950s to explain and predict health behaviours and used in practice to guide health promotion programmes[1,2]. It is a widely used model to assess personal beliefs and predict health behaviours, and it is based around the idea that people are more inclined to change their health behaviours if they believe that they are at risk. Figure 1 illustrates a summary of the Health Belief Model. As a consequence of the COVID-19 pandemic, there is an emerging stream of literature using the HBM to investigate factors determining people’s likelihood to get vaccinated against COVID-19[3–7]. To our knowledge we are the first to apply HBM constructs to the use of immunity certificates, as well as implement survey items informed by qualitative research thus proving new insights into determinants of immunity certificates public views.

**Figure 1.**
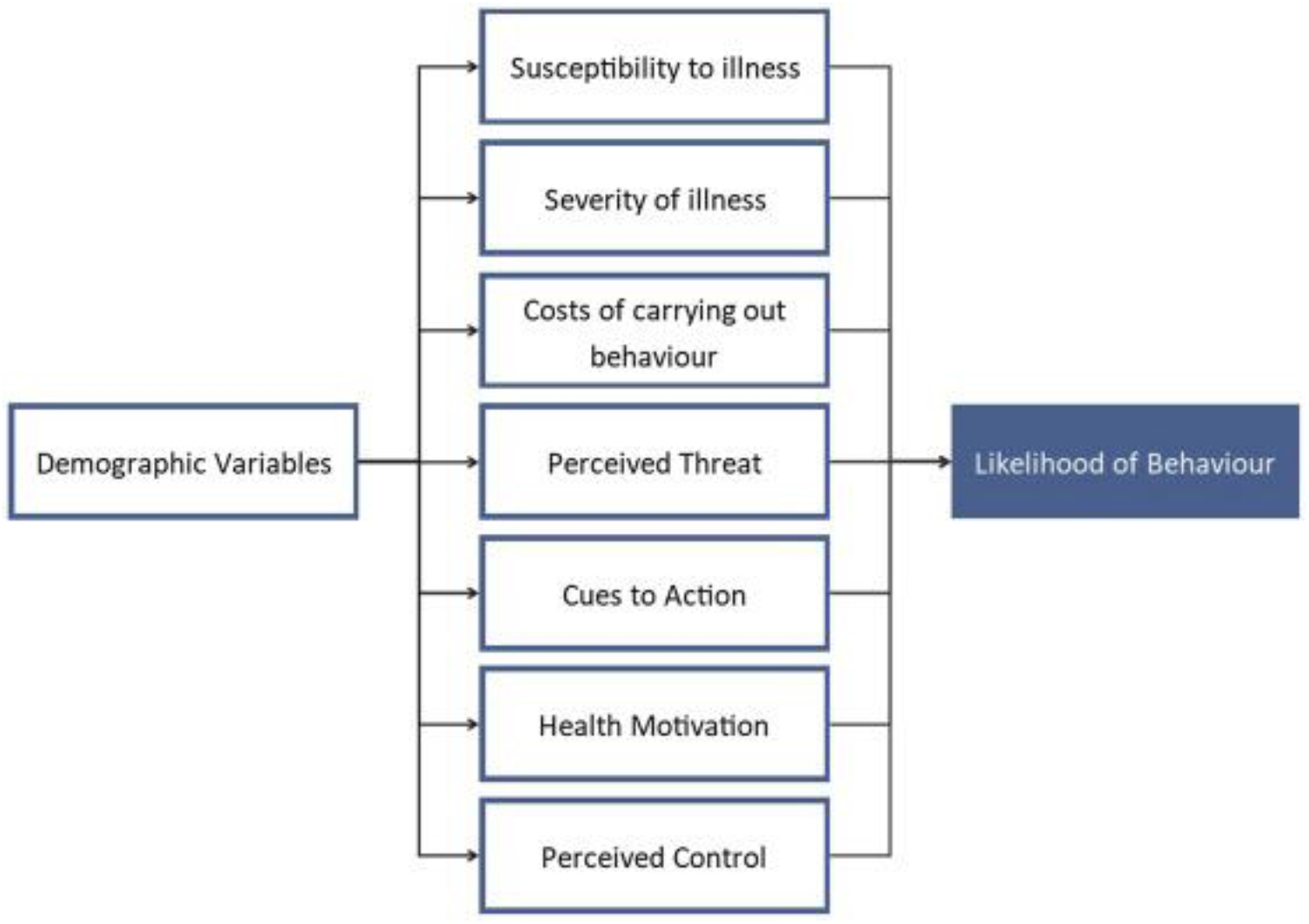
A schematic representation of the Health Belief Model sourced from Sciencedirect[8]

The HBM has six different constructs with multiple questions each: perceived susceptibility, perceived severity, perceived benefits of taking action, perceived barriers from taking action, cues to action, and self-efficacy in some models. In the cited literature these six dimensions refer to people’s perceptions on vaccination and COVID-19. However, immunity certification is a complex socio-technical system, and people’s views on it encompass beliefs around vaccination, COVID-19, and immunity certificates in general. Therefore, we adapted the HBM to fit the unique challenges of immunity certificates. The full questions, summary statistics and internal reliability of all HBM measures are reported in Table 1.

**Table 1.** Summary statistics of HBM measures

| HBM Measures | Items | Mean | Median | Std. Dev | Min | Max | Alpha |
| --- | --- | --- | --- | --- | --- | --- | --- |
| Perceived Severity of not using | I feel that without this service I won't be able to return to my workplace. | 2.4476 | 2 | 1.1558 | 1 | 5 | 0.8485 |
| Immunity Certificates | I feel that without this service my chances of getting a job will be affected. | 2.5918 | 3 | 1.1631 | 1 | 5 |  |
|  | I feel that without this service I won't be able to book face-to-face appointments with my GP/dentist. | 2.8371 | 3 | 1.2455 | 1 | 5 |  |
|  | I feel that without this service I won't be able to go to the theatre/movies/sports events. | 3.2715 | 4 | 1.1636 | 1 | 5 |  |
|  | I feel that without this service I won't be able to travel internationally. | 3.912 | 4 | 1.1252 | 1 | 5 |  |
|  | I feel that without this service I will not enjoy the same liberties I did before the pandemic. | 3.6667 | 4 | 1.1692 | 1 | 5 |  |
| Perceived COVID-19 Susceptibility | I am at risk of getting COVID-19 (SARS-CoV-2). | 3.5243 | 4 | 1.1255 | 1 | 5 | 0.7095 |
|  | It is likely that I will get COVID-19 (SARS-COV-2). | 2.9401 | 3 | 1.0122 | 1 | 5 |  |
|  | Individuals in my household are at risk for getting COVID-19 (SARS-COV-2). | 3.4438 | 4 | 1.131 | 1 | 5 |  |
|  | I feel knowledgeable about my risk of getting COVID-19 (SARS-COV-2). | 4.1255 | 4 | 0.746 | 1 | 5 |  |
| Perceived COVID-19 Severity | I believe that COVID-19 (SARS-COV-2) is a severe health problem in general. | 4.2266 | 4 | 0.9662 | 1 | 5 | 0.7061 |
|  | If I get COVID-19 (SARS-COV-2) I will get sick. | 3.7247 | 4 | 0.9749 | 1 | 5 |  |
|  | If I get COVID-19 (SARS-COV-2) I will die. | 2.1386 | 2 | 0.9227 | 1 | 5 |  |
|  | If I get COVID-19 (SARS-COV-2) other members in my household will get sick. | 3.5824 | 4 | 1.012 | 1 | 5 |  |
| Perceived Benefits of Immunity Certificates | This service will make me feel safe only if immunity is obtained through complete course of vaccination. | 3.2809 | 3 | 1.141 | 1 | 5 | 0.6045 |
|  | This service will make me feel safe only if immunity is obtained through past COVID-19 (SARS-CoV-2) infection. | 2.4326 | 2 | 0.9734 | 1 | 5 |  |
|  | This service will facilitate economic recovery. | 3.5506 | 4 | 1.054 | 1 | 5 |  |
|  | This service will facilitate social gatherings in closed spaces without restrictions (e.g. wearing masks, limits on number of people who can gather). | 3.7154 | 4 | 0.9922 | 1 | 5 |  |
| Perceived Barriers of using Immunity Certificates | I'm afraid that my data will be passed on to third parties without my consent or commercialized. | 3.0281 | 3 | 1.2883 | 1 | 5 | 0.3691 |
|  | This service will be difficult for me to use if available only on smartphones / tablets. | 1.9307 | 2 | 1.1913 | 1 | 5 |  |
|  | This service will be difficult for me to access if offered exclusively in English. | 1.2809 | 1 | 0.6982 | 1 | 5 |  |
| Vaccine Views | I am not convinced that the vaccine will protect me against COVID-19 (SARS-CoV-2). | 2.3034 | 2 | 1.2212 | 1 | 5 | 0.3276 |
|  | I feel worried about people who have received a non-UK approved vaccine entering the country. | 2.6292 | 3 | 1.2055 | 1 | 5 |  |
|  | I feel less at risk from catching coronavirus if I'm around people who have been fully vaccinated. | 3.8464 | 3 | 1.1257 | 1 | 5 |  |

Firstly, we adapt ***perceived COVID-19 susceptibility*** and ***perceived COVID-19 severity*** from two studies exploring people’s willingness to get vaccinated from COVID-19 using the HBM[4,5]. Each item is rated on a 5-point Likert scale ranging from 1 (“Strongly disagree”) to 5 (“Strongly agree”). We adapt the first three items for *COVID-19 Perceived Susceptibility* from [4] and the fourth item from [5] (Figure 1). Similarly, all four items measuring *COVID-19 Perceived Severity* are adapted from [4]. Cronbach’s alpha is 0.7061 for *COVID-19 Perceived Susceptibility* and 0.7095 for *COVID-19 Perceived Severity*, suggesting good internal consistency[9]. Therefore, these two constructs can be aggregated into two indices for statistical analysis purposes. We created two indices *COVID-19 Perceived Susceptibility* and *COVID-19 Perceived Severity* by averaging the four items within each construct[10,11].

Following that we created three items by adapting the HBM constructs for health behaviours to immunity certificates. As such we measure ***perceived benefits of using immunity certificates, perceived barriers from using immunity certificates***, and ***perceived severity of not using immunity certificates***. In the context of HBM and COVID-19 vaccination these items would usually refer to perceived benefits of vaccination, perceived barriers from getting vaccinated, and perceived severity of COVID-19. We kept the same questionnaire structure that is normally used for HBM studies and that we used to measure *COVID-19 Perceived Susceptibility* and *COVID-19 Perceived Severity*, but we formulated the questions around the use of immunity certificates.

The contents of the questions measuring ***perceived benefits of using immunity certificates, perceived barriers from using immunity certificates***, and ***perceived severity of not using immunity certificates*** were informed by our own findings from a series of qualitative studies including focus groups and interviews[12]. Using the findings of qualitative research to inform questionnaire survey question is a common practice[13]. At the time when this study was conducted literature on immunity certificates was still limited, hence other examples of research using HBM for immunity certificates were not available. For this reason, we used our own qualitative findings to inform the HBM survey items and generalize our findings to the general public.

Public views on immunity certificates from the perspective of users and experts in virology, public health, policymaking, bioethics, law, data science and artificial intelligence were gathered during the first focus groups in May 2021. Those results were then complemented by the perspective of service providers from different industries (culture, airlines, hospitality, and sports) obtained during the interviews between May and September 2021. We then used the themes that emerged from those findings to inform the design of survey questions regarding perceived severity of not using immunity certificates, perceived benefits of using immunity certificates, and perceived barriers of not using immunity certificates. Figure 2 presents some of the views on immunity certificates expressed during the focus groups conducted in May 2021 for illustration purposes. The full outcomes of the focus group research are aggregated in a map of the immunity certificate socio-technical system[14].

**Figure 2.**
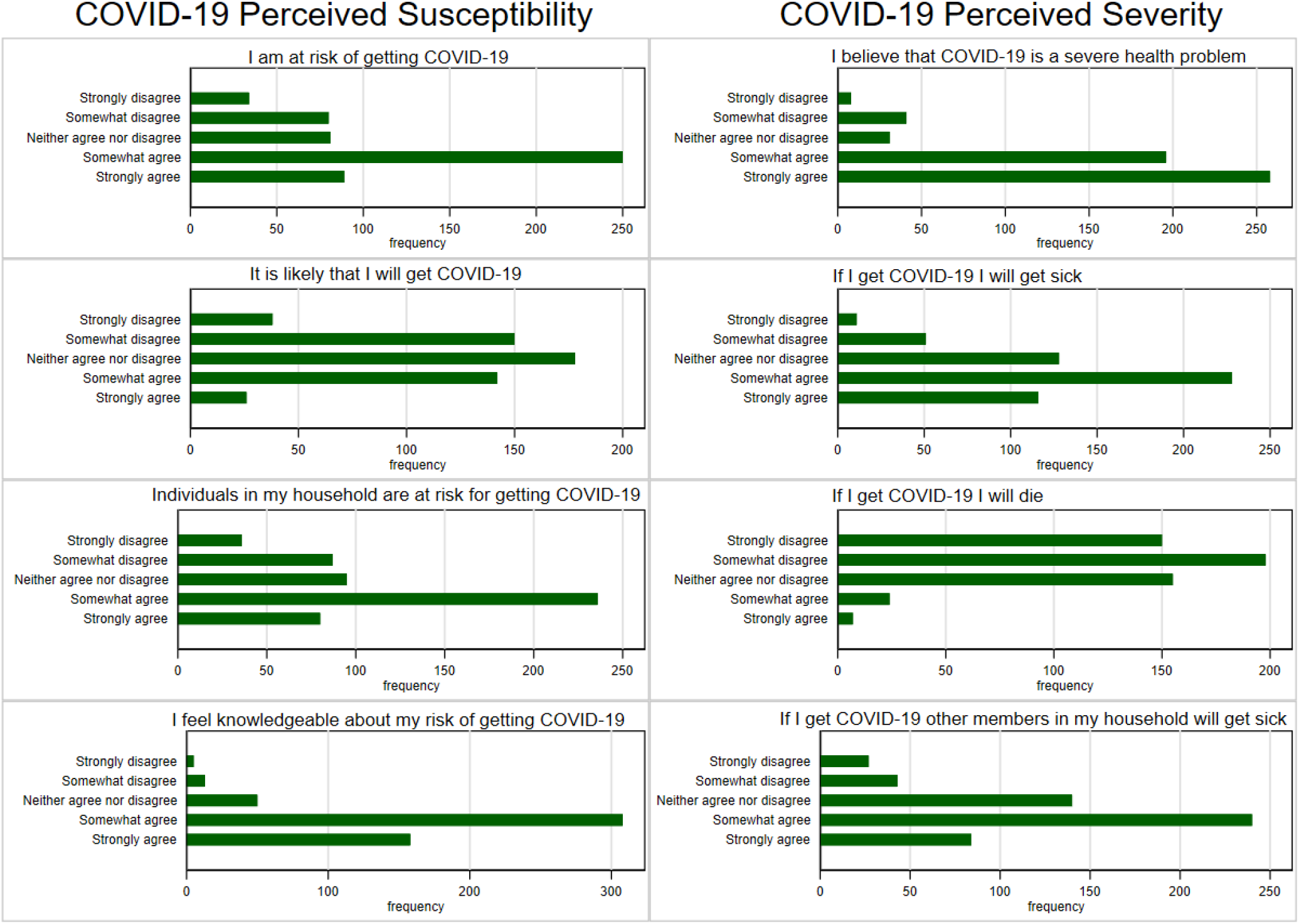
Distribution of answers by COVID-19 Perceived Susceptibility and COVID-19 Perceived Severity

The main three ***barriers from using immunity certificates*** that we identified were data safety, digital exclusion for those who do not have the means/capacity to use smartphones, and language barriers (Table 1, Figure 3).

**Figure 3.**
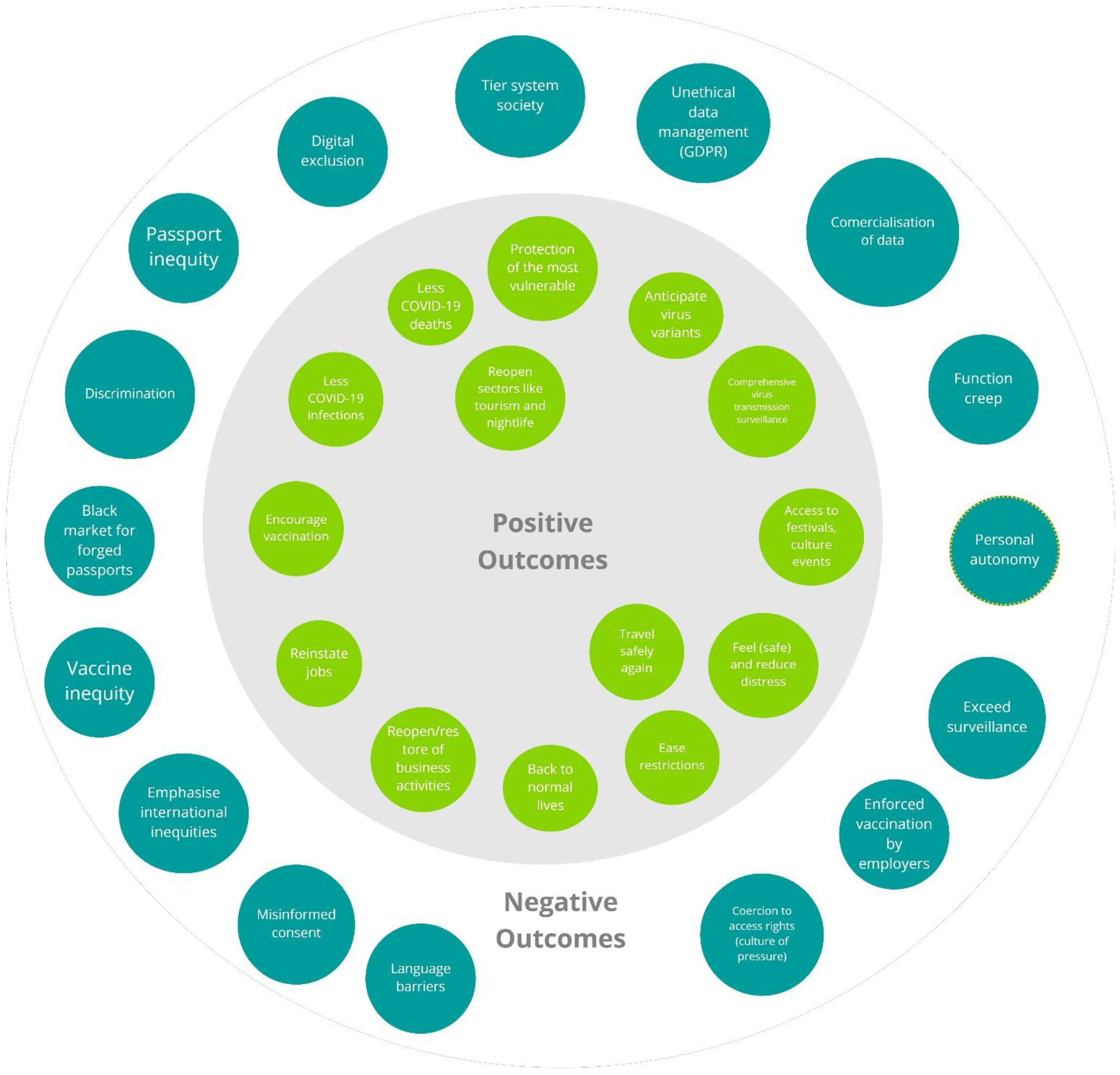
Sample of perceived outcomes of using immunity certificates: focus groups findings

Following that we identified four main ***perceived benefits of using immunity certificates***: (i) feeling safe if immunity was obtained through vaccination, (ii) feeling safe if immunity was obtained through past infection, (iii) economic recovery, and (iv) facilitating social gatherings (Table 1, Figure 4). We split the benefit of feeling safe into two different questions addressing immunity through vaccination or past infection. One reason for this differentiation was because during our focus groups participants voiced confusion regarding the official definition of COVID-19 immunity. Another reason was that some participants voiced concerns regarding the efficacy and duration of immunity obtained through past infection.

**Figure 4.**
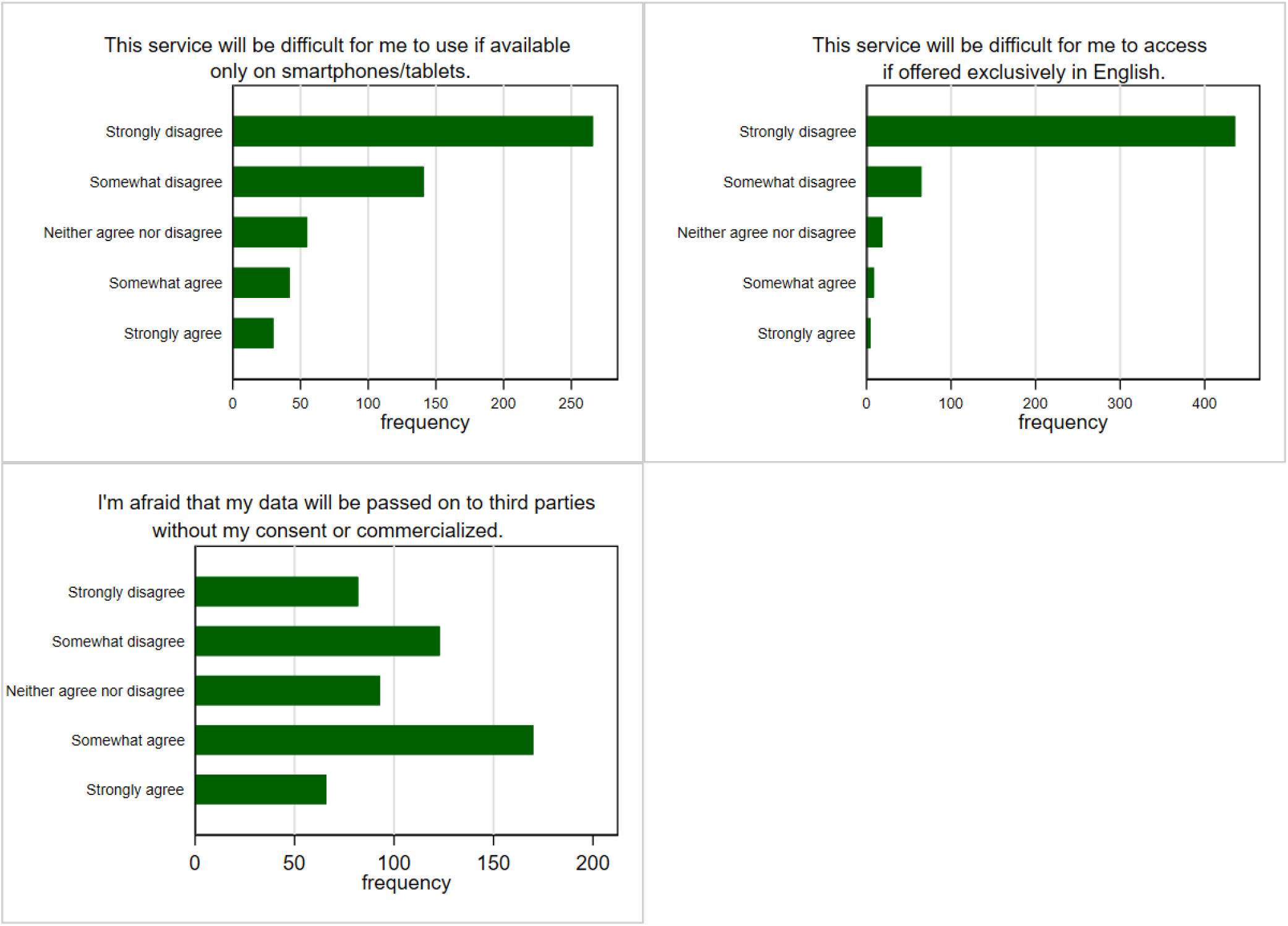
Distribution of answers by perceived barriers from using immunity certificates from the cross-sectional survey *(https://osf.io/jubv6/)*

The perceived severity of not using immunity certificates in our study is a similar concept to ***perceived severity of COVID-19*** and perceived severity of COVID-19 vaccines in traditional HBM research, which was adapted to fit a service rather than a virus or disease. We created six questions to measure this concept based on a combination of unintended consequences, positive and negative outcomes of immunity certificates voiced by participants in previous focus gr oups. Therefore, the questions refer to returning to work, travelling, attending different social or cultural events, and enjoying the same pre-pandemic liberties (Table 1, Figure 5). Cronbach’s alpha for perceived severity of not using immunity certificates was 0.8485 showing good internal consistency (from the data in our cross-sectional study). Therefore, the six items were also aggregated into an index of ***perceived severity of not using immunity certificates***, by taking the average score of all items as done previously.

**Figure 5.**
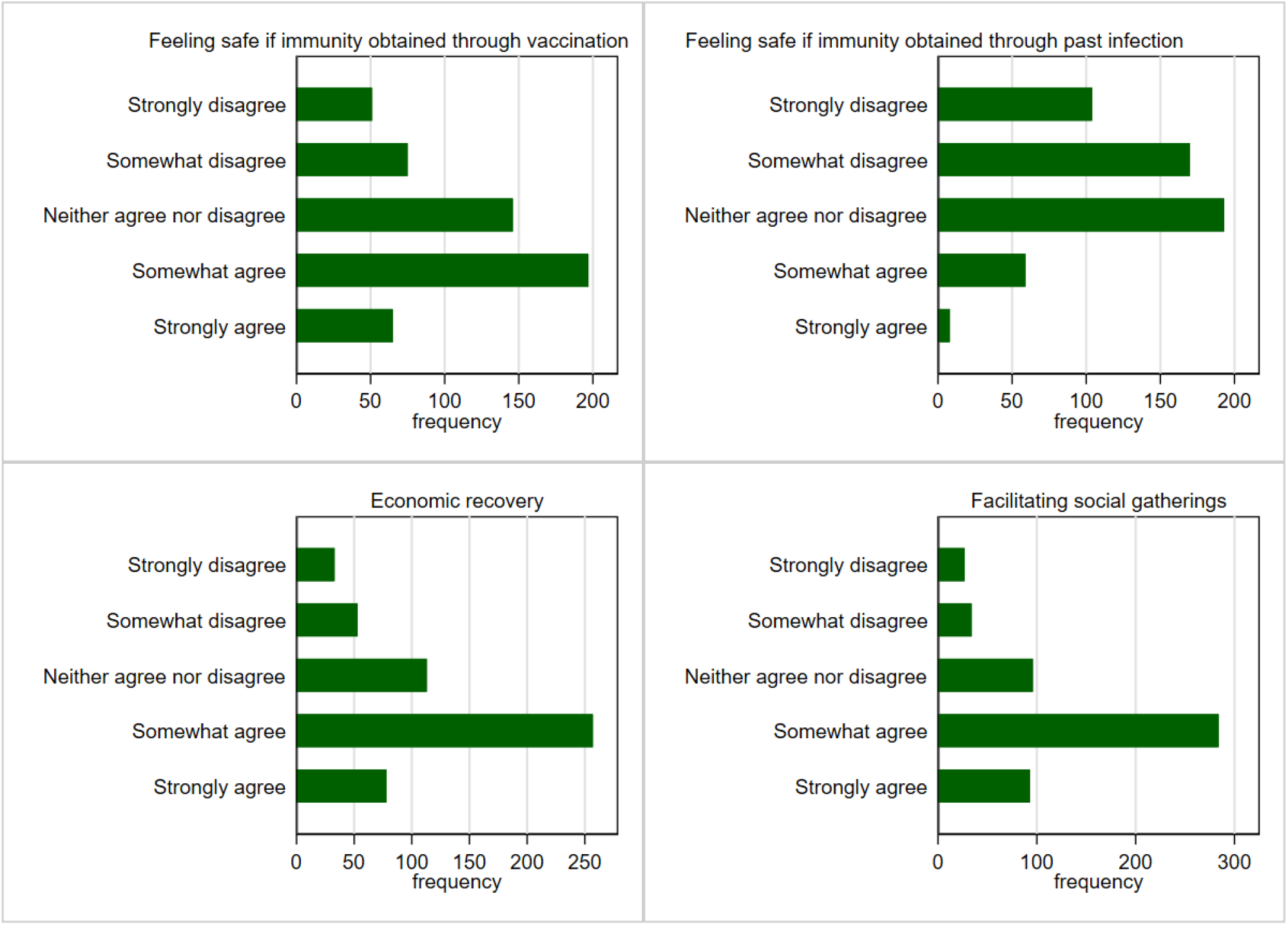
Distribution of answers by perceived benefits of using immunity certificates from the cross-sectional survey *(https://osf.io/jubv6/)*

Finally, as the use of immunity certification is partly dependent on ***COVID-19 vaccinations*** we construct a series of question on this topic. However, we did not employ the conventional HBM constructs measuring the public’s intention to get vaccinated, vaccination barriers or perceived severity of COVID-19 vaccines. The reason for this was that the time when the present study was conducted approximately 75% of the UK’s adult population had been vaccinated[15]. Therefore, intention to get vaccinated or barriers from vaccinations were not as relevant anymore, at least for UK-based studies like ours. As such we constructed three questions on COVID-19 vaccination based on our qualitative research findings. The three questions were (i) worries that the vaccine is not effective, (ii) worries about non-UK approved vaccines, and (iii) feeling safe around vaccinated people (Table 1, Figure 6).

**Figure 6.**
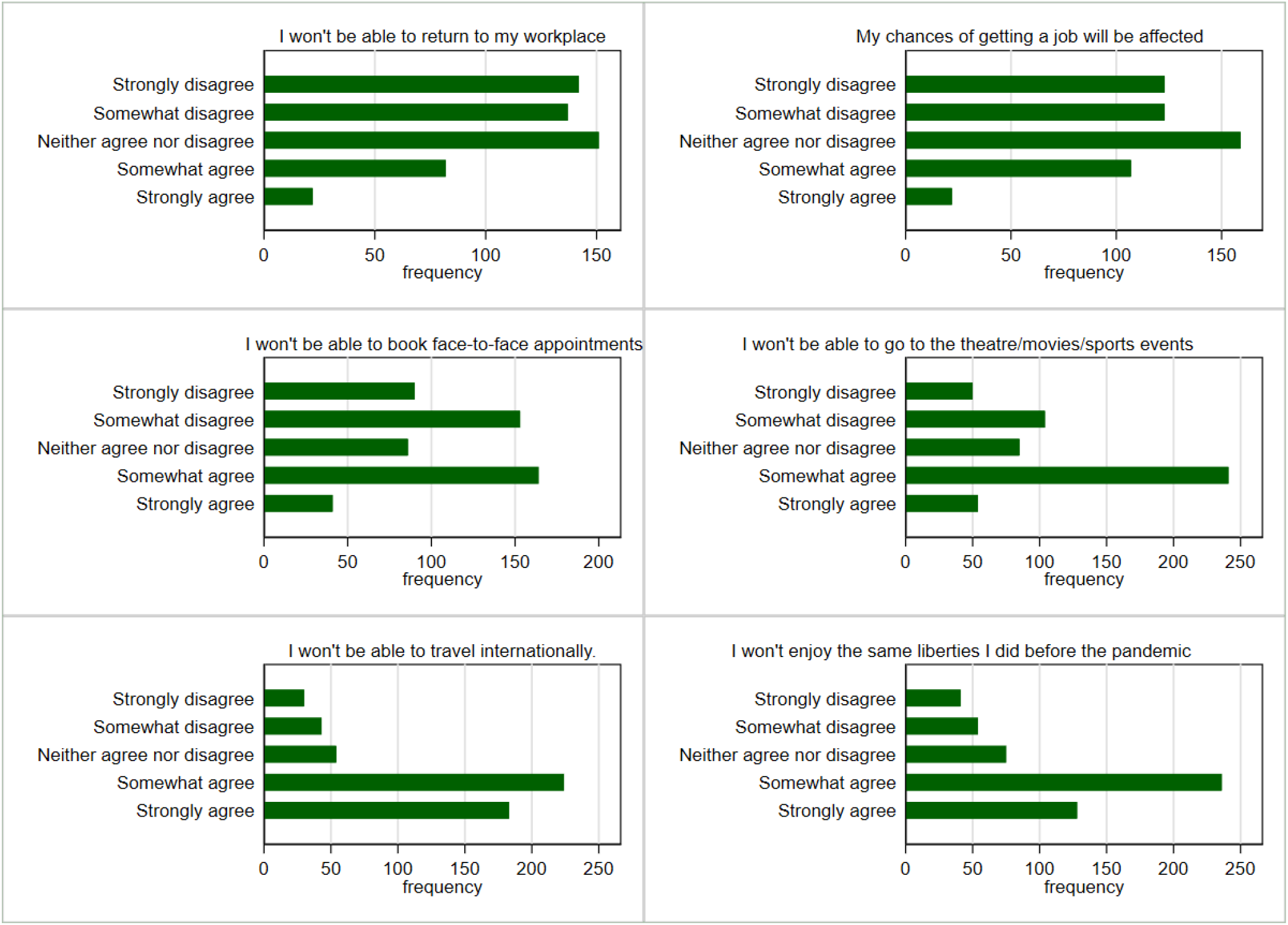
Distribution of answers by perceived severity of not using immunity certificates from the cross-sectional survey *(https://osf.io/jubv6/)*

**Figure 7.**
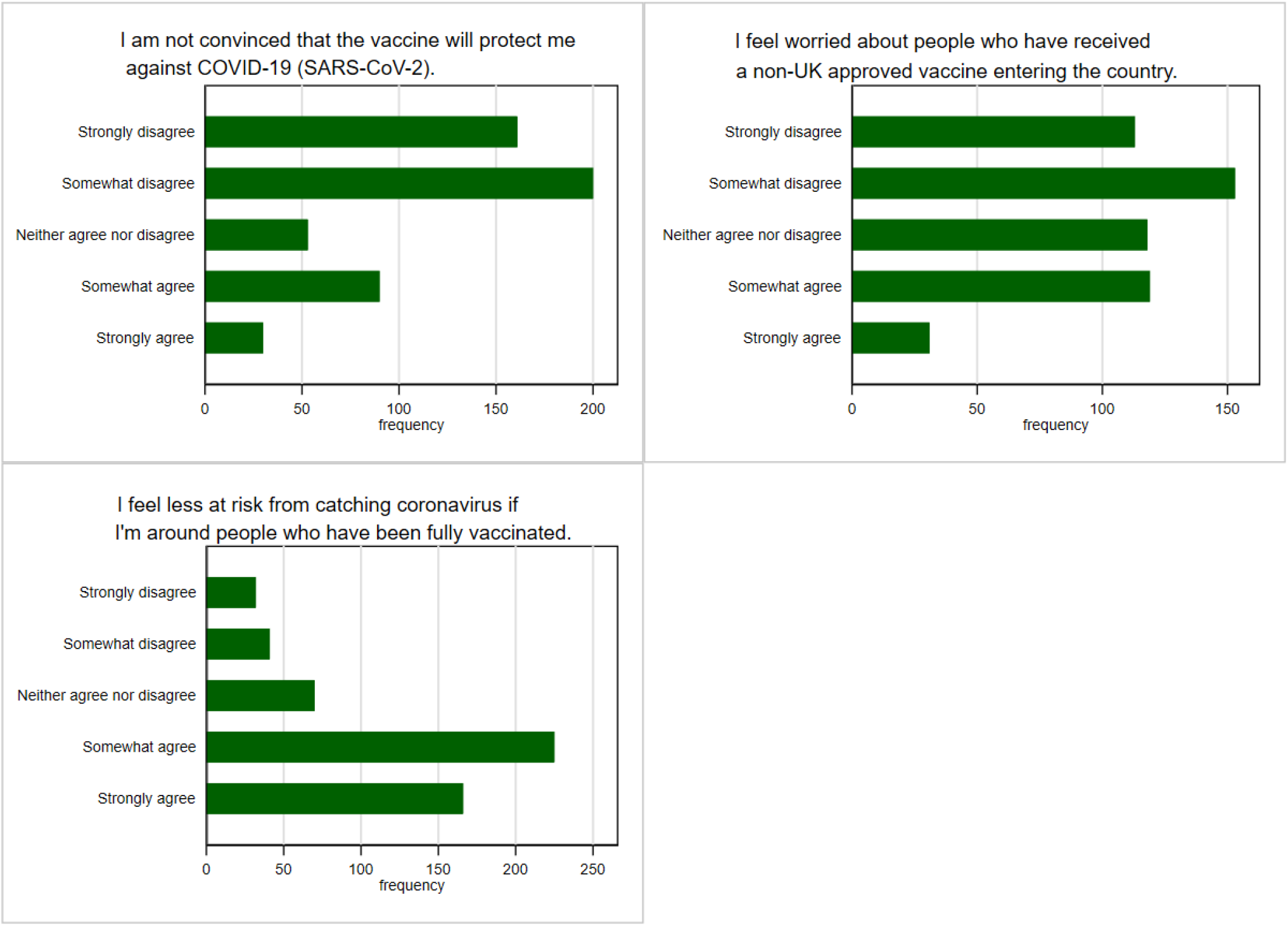
Distribution of answers by COVID-19 vaccination views from the cross-sectional survey *(https://osf.io/jubv6/)*

The survey items comprising the perceived barriers and perceived benefits of using immunity certificates displayed a low Cronbach’s alpha of 0.3691, 0.6045 and respectively 0.3276, hence these items were not aggregated into individual indices, but rather used on their own. Our analysis was an exploratory one, and the main of the survey was to explore willingness to use immunity certificates in different scenarios[16]. Further research should take a theoretical approach in creating survey items for applying the HBM to immunity certificates.

## Data Availability

The data is available in a public, open access repository. All statistical code, dataset, survey item and ethical approval are available on OSF (https://osf.io/jubv6/).

## Footnotes

### Contributors

The questionnaire survey was conceptualised by CN, IS and PB, with the input of TG, CLA, and OC. CEN and PB completed the data collection. CN and IS conducted the statistical analysis. All authors contributed and approved the final manuscript.

### Ethics statements

Ethics approval was obtained from the College of Engineering, Design and Physical Sciences Research Ethics Committee at Brunel University London (Ref. 31705-A-Jul/2021-33586-1) on the 29^th^ of July 2021.Informed consent was obtained from all respondents prior to the beginning of the survey. Respondents were allowed to withdraw from the survey at any time.

